# Will ChatGPT-4 improve the quality of medical abstracts?

**DOI:** 10.1101/2024.02.09.24302591

**Authors:** Jocelyn Gravel, Chloé Dion, Mandana Fadaei Kermani, Sarah Mousseau, Esli Osmanlliu

**Affiliations:** Department of Pediatric Emergency Medicine, CHU Sainte-Justine, Université de Montréal, Montréal, Québec, Canada; Faculté de médecine, Université de Montréal, Montréal, Québec, Canada; Division of Pediatric Emergency Medicine, Montreal Children’s Hospital, McGill University, Montréal, Québec, Canada

**Author notes:** **Corresponding author:** Jocelyn Gravel, Department of Pediatric Emergency Medicine, CHU Sainte-Justine, 3175 Chemin de la Côte-Sainte Catherine, Room A326 Montreal (Quebec) H3T 1C5 Canada, #2559.

**Keywords:** ChatGPT, Medical informatics, Editing

## Abstract

**Background:** ChatGPT received recognition for medical writing. Our objective was to evaluate whether ChatGPT 4.0 could improve the quality of abstracts submitted to a medical conference by clinical researchers.

**Methods:** This was an experimental study involving 24 international researchers who provided one original abstract intended for submission at the 2024 Pediatric Academic Society (PAS) conference. We created a prompt asking ChatGPT-4 to improve the quality of the abstract while adhering PAS submission guidelines. Researchers received the revised version and were tasked with creating a final abstract. The quality of each version (original, ChatGPT and final) was evaluated by the researchers themselves using a numeric scale (0-100). Additionally, three co-investigators assessed abstracts blinded to the version. The primary analysis focused on the mean difference in scores between the final and original abstracts.

**Results:** Abstract quality varied between the three versions with mean scores of 82, 65 and 90 for the original, ChatGPT and final versions, respectively. Overall, the final version displayed significantly improved quality compared to the original (mean difference 8.0 points; 95% CI: 5.6-10.3). Independent ratings by the co-investigator confirmed statistical improvements (mean difference 1.10 points; 95% CI: 0.54-1.66). Researchers identified minor (n=10) and major (n=3) factual errors in ChatGPT’s abstracts.

**Conclusion:** While ChatGPT 4.0 does not produce abstracts of better quality then the one crafted by researchers, it serves as a valuable tool for researchers to enhance the quality of their own abstracts. The utilization of such tools is a potential strategy for researchers seeking to improve their abstracts.

**Funding:** None

## Introduction

Natural Language Processing (NLP) is a field of artificial intelligence that focuses on the interaction between computers and humans using natural language(1). Large language models (LLM) refer to a breakthrough in the field of NLP, distinguished by their large size and pre-training on extensive datasets to acquire a broad understanding of language patterns(2). ChatGPT is a LLM-enabled chatbot able to produce text in response to multiple types of inputs (3). Since its launch in November 2022, scientific articles partially written by ChatGPT have been published(4-6). While many authors have acknowledged the quality of the writing(7-13) many journals questioned the ethical aspects of using chatbots for scientific writing(14-18). Researchers identified important limitations of using ChatGPT for writing including many demonstrations that it commonly provides inaccurate references(19-21).

Many articles reported that ChatGPT can write credible abstracts(22, 23), but few studies evaluated the ability of ChatGPT to write scientific abstracts. Gao et al. compared 50 abstracts from real publications to abstracts generated by ChatGPT and concluded that “*ChatGPT writes believable scientific abstracts*”(24). On the other hand, Ali and Singh advised that abstracts written by ChatGPT must be verified to detect self-additions(25).

To our knowledge, no study has quantitatively evaluated the quality of abstracts provided by ChatGPT and its potential for helping researchers improve their work. Our main objective was to evaluate whether ChatGPT could improve the quality of abstracts submitted to conferences by clinical researchers.

## Methods

This was an experimental study conducted in October 2023 evaluating the benefit of asking ChatGPT (Version 4.0 OpenAI Inc, San Francisco, CA, USA) to revise abstracts submitted to a medical scientific conference.

A convenience sample of researchers was invited to provide one abstract that they expected to submit to the 2024 Pediatric Academic Societies (PAS) conference. The researchers were identified by two co-investigators based on their experience of publication and previous participation to the PAS conference.

The intervention of interest was the use of ChatGPT-4 to improve the abstract. To do this, we created the following prompt asking ChatGPT-4 to improve the quality of the abstract while adhering PAS submission guidelines: “*Using the following guidelines: 1. Have a title; 2. Have an abstract containing the following five sections: introduction, objective, method, results and conclusion; have a maximum of 2600 characters (with space excluded title), improve the following scientific abstract for clarity:* **insert abstract**”. Each abstract constructed by ChatGPT was returned to its his corresponding researcher within 24 hours following reception of the original abstract. Researchers were then invited to provide a final abstract using nothing, parts or the entire ChatGPT version to improve their abstract. It was expected that the final version would be completed following reception of the ChatGPT version. In the end there were three versions of each abstract:

### 1. Original version

The first version of the abstract as initially submitted by the researcher.

### 2. ChatGPT version

The abstract constructed by ChatGPT using the prompt asking to improve the original version.

### 3. Final version

The version that the researcher used at the end of the process.

The primary outcome was the quality of the abstract measured with a verbal numeric scale from 0 to 100, using the following instruction: *On a scale of 0 to 100% on the quality of the abstract, where 0% means poor quality and 100% the best possible abstract, how would you rate the abstract?* This evaluation was initially conducted by the invited researchers considering they are the experts in the field of their abstract and because our primary outcome was to evaluate if ChatGPT could help researchers. In that way, their opinion is the most important. Multiple secondary outcomes were measured. Among them, the quality of the original and final versions of the abstracts were evaluated by three members of the research team using the same verbal numeric scale. This evaluation was blinded to the version (original vs final). Moreover, researchers identified whether the abstract fulfilled all requirements of the PAS conference, and the presence of any factual error. These errors were classified as minor (no impact on the conclusion of the abstract or minimal impact on the probability of acceptance of the abstract by the conference reviewers) or major. Researchers also reported whether they consider that the use of ChatGPT improved the quality of their abstract (yes/no), and if it improved their chance of being accepted (yes/no). Researchers were asked if the ChatGPT version could have been submitted as is. Reasons justifying the last answer were collected.

Few independent variables were collected as potential factors associated to the outcomes. These were related to the writer of the abstract (number of years as researcher, number of abstracts previously written, previous use of ChatGPT, fluidity in English) and the abstract (qualitative vs quantitative study, presence of a table or a figure).

The primary analysis was the mean difference in scores between the final and the original version of the abstract according to the participating researchers. To do this, we calculated the difference (final abstract score minus original abstract score) for each abstract and report the mean and 95% confidence interval (95% CI) for these differences. Other analyses included the mean score for each version (original, ChatGPT, and final) and differences according to the score assigned by the three co-authors.

The proportion of researchers responding that the use of ChatGPT improved their abstract, and the number of abstracts in which a factual error was found were calculated. Finally, researchers reported their final evaluation of the usefulness of the revision provided by ChatGPT.

An exploratory analysis was carried out to evaluate factors associated with a positive impact of the use of ChatGPT. To do this, we evaluated the association between independent variables related to the researchers (age group, gender, experience with abstract submission and ChatGPT) and to the abstracts (type, presence of figure), and improvement in score using ANOVA.

We had no prespecified idea of the mean scores for the evaluation of the abstracts. However, it was expected that the final abstract should not have a lower score than the original one as they are both written and evaluated by the same researcher who should improve or keep the same abstract. Based on our previous experience, we anticipated small but constant improvement of the quality of abstracts using ChatGPT. Aiming for an effect size of 0.7, it was estimated that we needed to evaluate at least 20 abstracts.

Our local Institutional Review Board concluded that no ethical review was required for the project as researchers are viewed as raters and not participants. Consequently, providing an abstract and responding to our invitation to review was considered as a consent to participate in the study.

## Results

A total of 33 researchers were directly invited to collaborate in the study. Among them, six did not plan to submit an abstract, one did not respond, and one was not at ease to use ChatGPT for abstract generation. In the end, 25 researchers provided an abstract and 24 (96%) completed the evaluation of their three abstracts.

Table 1 provides a comprehensive overview of the researchers and abstracts engaged in the study. In brief, 15 studies (63%) employed quantitative methodologies, and 7 studies (30%) incorporated tables or figures to enhance data representation. The research team exhibited notable diversity, with 50% of participants being under the age of 34, 58% identifying as female, and 50% boasting a track record of more than 15 published abstracts. Regarding ChatGPT usage, 42% of participants had never utilized the tool, 50% had used it sporadically, and 8% employed it regularly. Proficiency in English was evenly distributed across various comfort levels, with 8 (33%) participants reporting their proficiency as moderately, quite, or very comfortable respectively.

**Table 1.** Characteristics of the abstracts and participants (n=24)

|  | N (%) |
| --- | --- |
| Type of study<br>Quantitative<br>Qualitative<br>Other | 15 (63)<br>5 (21)<br>4 (17) |
| Presence of table/figure | 7 (30) |
| Researchers' information |  |
| Age group<br>18-24 years<br>25-34 years<br>35-44 years<br>45-54 years<br>55-64 years | 3 (13)<br>9 (38)<br>7 (30)<br>4 (17)<br>1 (4) |
| Gender<br>Female<br>Male | 14 (58)<br>10 (42) |
| Number of previous abstracts<br>This is the first<br>1-5<br>6-15<br>> 15 | 1 (4)<br>7 (30)<br>4 (17)<br>12 (50) |
| ChatGPT previous experience<br>Never used it<br>Used it a few times<br>Used it many times | 10 (42)<br>12 (50)<br>2(8) |
| English literacy<br>Moderately comfortable<br>Quite comfortable<br>Very comfortable | 8 (33)<br>8 (33)<br>8 (33) |

Abstract quality varied between the three versions with mean scores of 82 (95%CI: 78-86), 65 (95%CI: 58-73) and 90 (95%CI: 88-93) for the original, ChatGPT and final versions, respectively. Overall, the final abstracts displayed significantly improved scores compared to the original ones according to the researchers with improvement varying between 1 and 20 points (mean difference 8.0 points; 95% CI: 5.6-10.3). Of note, only three (14%) abstracts scores remained unchanged in the final version. Figure 1 shows that for most series of abstract, the ChatGPT version had the lowest score, but the final version had a score higher than the original.

**Figure 1.**
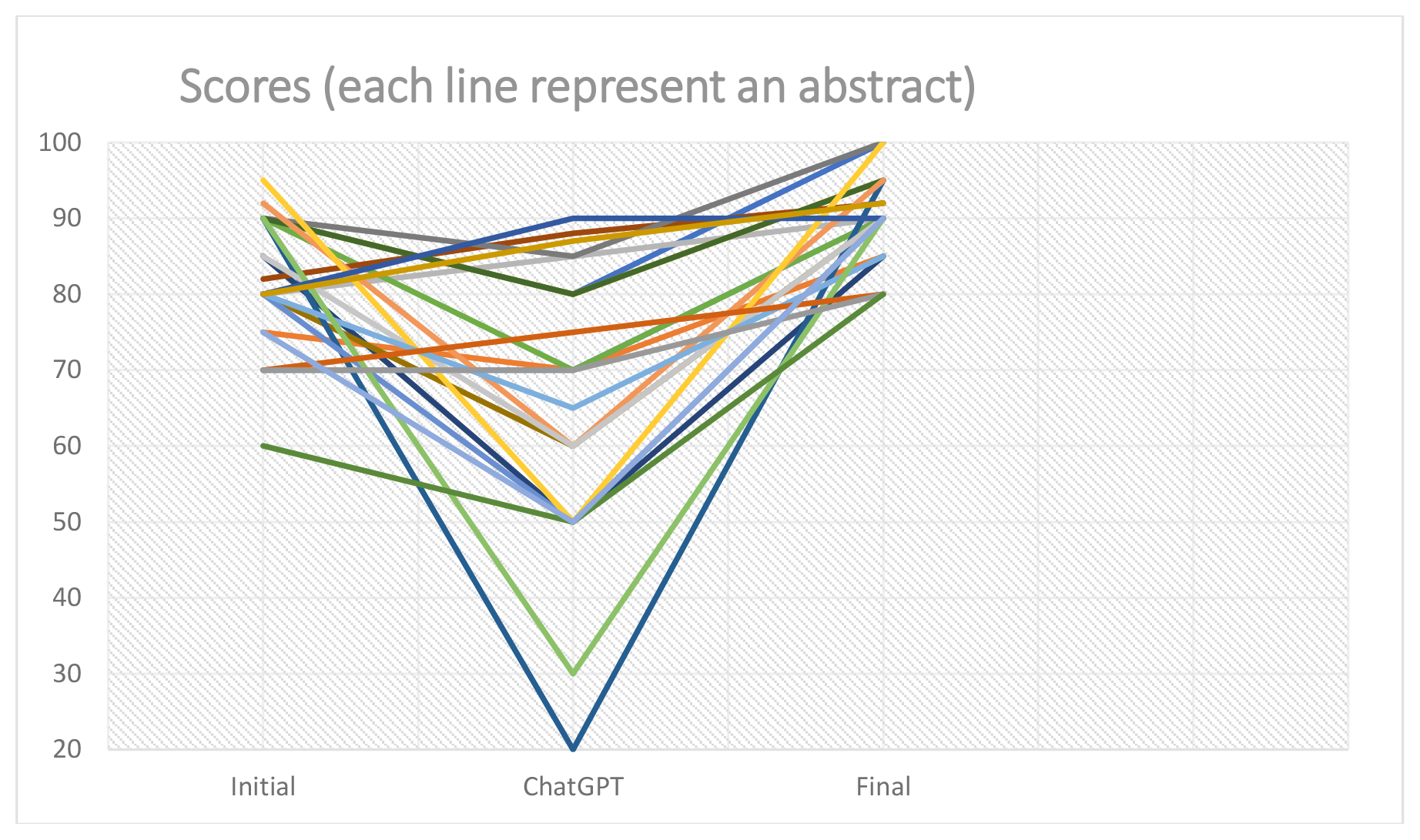
Scores of the initial, ChatGPT and final version of the abstracts

Researchers identified minor (n=10) and major (n=3) factual errors in the abstracts generated by ChatGPT. Examples of major errors included omission of providing the primary outcome in the method section or confusion/ omission in the results provided. Among the minor errors, ChatGPT omitted to provide confidence intervals in many abstracts. Eighteen (75%) participants reported that ChatGPT contributed to the enhancement of their final abstract and ten (42%) participants believed it improved their abstract’s probability of acceptance. Finally, 18 (75%) researchers reported being uncomfortable using the ChatGPT version for conference submission; mostly because of the omission of important information.

All 24 pairs of abstracts (original and final) were evaluated in triplicate by co-authors. Using the mean score of the three raters, there was a statistical improvement in scores for the final vs. the original version (mean difference 1.10 points; 95% CI: 0.54-1.66).

None of the independent variables were statistically associated to the improvement in scores (table 2). However, there was a significant pattern toward a higher impact of ChatGPT for researchers being less comfortable in English (p: 0.062) and for those who had less experience in abstract submission (0.085).

**Table 2.** Association between independent variables and improvement in score.

|  | Mean difference | p-value on ANOVA |
| --- | --- | --- |
| Type of study<br>Quantitative<br>Qualitative<br>Other | 6.6<br>11.0<br>9.3 | 0.282 |
| Presence of table/figure<br>No table/figure | 7.3<br>8.2 | 0.712 |
| Researchers' information |  |  |
| Age group<br>18-24 years<br>25-34 years<br>35-44 years<br>45-54 years<br>55-64 years | 11.7<br>7.9<br>6.7<br>7.0<br>10.0 | 0.779 |
| Gender<br>Female<br>Male | 8.9<br>6.7 | 0.360 |
| Number of previous abstracts<br>This is the first<br>1-5<br>6-15<br>> 15 | 20<br>9.43<br>7.5<br>6.3 | 0.085 |
| ChatGPT previous experience<br>Never used it<br>Used it a few times<br>Used it many times | 7.5<br>8.0<br>10.0 | 0.856 |
| English literacy<br>Moderately comfortable<br>Quite comfortable<br>Very comfortable | 11.4<br>7.5<br>5.0 | 0.062 |

## Discussion

This experimental prospective study demonstrated that the use of ChatGPT-4 led to a significant improvement in abstract quality, as reported by 24 international researchers. The mean difference of 8.0 points in abstract scores between the final and original versions suggests a substantial enhancement, supporting our hypothesis that ChatGPT-4 could contribute positively to the refinement of scientific abstracts.

To our knowledge, this is the first study evaluating the impact of the use of artificial intelligence to improve abstract writing. As mentioned, Gao et al. demonstrated that human can difficultly discriminate real abstracts from those generated by ChatGPT(24). However, they did not compare the quality of the abstracts. On the other hand, Khlaif et al. conducted an experimental study asking ChatGPT to produce four articles and 50 abstracts. They did not provide results related to the quality of the abstracts but concluded that ChatGPT can “improve the quality of high-impact research articles” (26).

The observed improvement in abstract quality seemed consistent across various demographic and experience-related variables. However, while none of these factors reached statistical significance, there was a trend toward a higher impact of ChatGPT for researchers who were less comfortable in English and those with less experience in abstract submission. This results aligns with Del Giglio et al. who suggested that artificial intelligence could improve scientific writing of non-native English-speaking scientists (27). It may also be helpful for less experienced researchers.

While the ChatGPT abstracts tended to have the lowest score, the final abstracts consistently surpassed the originals, suggesting that researchers used some parts of the ChatGPT version to improve their final version. The use of ChatGPT, could be seen as an external revision. The researchers’ discomfort with submitting the ChatGPT-generated abstracts for conference consideration is also a notable finding. Seventy-five percent of participants reported feeling uncomfortable using the ChatGPT version for submission, primarily due to omissions of critical information. This suggests that, despite the initial output, researchers played a crucial role in refining and improving the abstracts, leveraging ChatGPT as a valuable tool in the revision process.

The study must be put in the context of its limitations. Foremost among these limitations is the unblinded nature of the primary outcome measurement. This deliberate choice was motivated by our desire to directly ask researchers whether ChatGPT had a discernible positive impact and to quantify such perceived benefits. The subsequent blinded evaluation conducted by the three co-authors robustly validated the enhancements observed in the final version. Even though 75% of the researchers reported that the use of ChatGPT improved their final abstract, we were not able to measure the influence of external factors to improve the final abstract. For example, it is possible that just the fact of waiting 24 hours before revising the abstract had an impact on final version. We did not measure the acceptance rate of the abstracts. Ideally, we would have submitted both versions to the conference website and compare acceptance rates, but such a comparative analysis was deemed ethically untenable. Finally, this was a convenience sample, and it is possible that participants may have overestimated the impact of ChatGPT.

## Conclusion

In conclusion, our study demonstrates evidence supporting the potential of ChatGPT in enhancing the quality of scientific abstracts. While the tool has shown promise in helping researchers, vigilance is crucial to address factual errors and identify omission. As artificial intelligence continues to evolve, understanding its role in the scientific writing process becomes increasingly important.

## Data Availability

All data produced in the present study are available upon reasonable request to the authors

## Abbreviations

(ANOVA): Analysis of Variance
(CI): Confidence Interval
(LLM): Large Language Models
(NLP): Natural Language Processing
(PAS): Pediatric Academic Society

## Acknowledgements

The study team will acknowledge the contribution of all researchers who agreed to provide abstract and rate the quality of the responses for this study. This includes but is not limited to: Waleed Alqurashi, Naila Bouadi, Adalet Bugra, Brett Burstein, Mathieu Dehaes, Evelyne D. Trottier, Lea Dikranian, Olivier Drouin, Gabrielle Freire, Nathalie Gaucher, Borja Gomez, Nour Kabbes, Nirupama Kannikeswaran, Arielle Levy, Thuy Mai Luu, Keon Ma, Santiago Mintegi, Ahmed Moussa, Raphaelle Pelc, Soha Rached-D’Astous, Asa Rahimi, Christina Santamaria, Johan N. Siebert, Madeleine Sumner, Philippe Sylvestre, Sevag Tachejian.

## Declaration of generative AI and AI-assisted technologies in the writing process

During the preparation of this work the author(s) used ChatGPT 4.0 in order to improve language and readability. After using this tool, the author(s) reviewed and edited the content as needed and take(s) full responsibility for the content of the publication.

